# LLM-Driven Extraction of NI-RADS and Imaging Tumor Characteristics to Enhance Oropharyngeal Cancer Survivorship Surveillance

**DOI:** 10.64898/2026.06.11.26355483

**Authors:** Wenye Song, Lina Shbita, Isabelle Jia Hui Jang, Olga Starostina, Ryan Lewis, Ariana Sahli, Warren Floyd, Md Mahin, Waree Rinsurongkawong, Carly Barbon, Stephen Lai, J. Jack Lee, Komal Shah, Melissa Chen, Katherine A. Hutcheson, Clifton David Fuller, Amy Moreno, the OPC-SURVIVOR Research Program

**Author notes:** **Corresponding Author Amy C. Moreno, MD MS**, Department of Radiation Oncology, The University of Texas MD Anderson Cancer Center, Houston, TX, USA.

## Abstract

**Purpose:** Radiologic surveillance is essential for oropharyngeal cancer (OPC) survivors, guiding recurrence detection and follow-up strategies. The Neck Imaging Reporting and Data System provides a standardized framework for post-treatment risk reporting at both the primary tumor site (pNI-RADs) and cervical lymph nodes (nNI-RADS). Comprehensive surveillance additionally requires assessment of disease status, including the primary tumor, nodal involvement, and distant metastases. These clinical results are often embedded as unstructured data within free-text radiology reports. We hypothesized that a large language model (LLM) can reliably extract NI-RADS score criteria and summarize key imaging features from unstructured radiology text, achieving high concordance with expert review.

**Methods:** Previously untreated OPC patients who received definitive cancer therapy were identified. Eligible imaging reports included post-treatment head and neck CT, MRI, or FDG PET/CT scans containing narrative and impression text. Examinations lacking narrative or impression text, containing pre-existing NI-RADS annotations, or involving non-surveillance imaging modalities were excluded. A total of 200 reports were randomly selected from 7,076 eligible examinations for manual abstraction using a three-reviewer consensus framework to establish a reference dataset. Using the Palantir Foundry Pipeline Builder, a GPT-5-based LLM was deployed to extract pNI-RADS and nNI-RADS scores, and key imaging features of disease status from these reports. Performance was evaluated using exact agreement and F1-based metrics.

**Results:** Agreement for no evidence of disease (score of 1) was 93.3% (126/135; F1 = 0.94) and 90.3% (130/144; F1 = 0.93) for pNI-RADS and nNI-RADS, respectively. For NI-RADS ≥2, exact category agreement was 73.1% (38/52; macro-F1 = 0.75) for pNI-RADS and 64.3% (27/42; macro-F1 = 0.56) for nNI-RADS. Quadratic weighted κ was 0.81 and 0.59, respectively. For post-treatment disease surveillance variables, agreement was 94.9% (149/157; F1 = 0.87) for primary tumor presence, 89.1% (164/184; F1 = 0.87) for nodal disease presence, and 94.7% (126/133; F1 = 0.70) for distant metastasis detection. Specificity was high across disease-status variables (0.95-0.99), with negative predictive values of 0.95 for primary tumor, 0.87 for nodal disease, and 0.99 for distant metastasis.

**Conclusions:** Our LLM-based information retrieval and classification approach for radiographic treatment response from unstructured, multidimensional imaging reports achieved high performance for disease exclusion and moderate performance for detecting suspected residual and/or new disease. This pipeline supports scalable and standardized surveillance data capture for longitudinal monitoring, clinical analytics, and survivorship research in head and neck oncology.

## Introduction

Oropharyngeal cancer (OPC) survivorship is rapidly expanding, driven largely by the increasing incidence of human papillomavirus (HPV)-associated disease and substantial improvements in long-term survival following multimodality treatment^1,2^. As a result, a growing population of OPC survivors now requires prolonged post-treatment surveillance to monitor for locoregional recurrence, nodal relapse, and distant metastatic progression. Early identification of recurrent disease remains clinically important because salvage interventions are often most effective when recurrence is detected at a limited stage, directly influencing treatment options, prognosis, and long-term functional outcomes. In addition to recurrence detection, surveillance imaging also provides critical information regarding treatment response, residual abnormalities, and evolving disease burden over time^3,4^.

The Neck Imaging Reporting and Data System (NI-RADS) was developed to standardize interpretation and reporting of post-treatment head and neck imaging, providing a structured framework for recurrence risk stratification at both the primary tumor site (pNI-RADS) and cervical lymph nodes (nNI-RADS)^5^. NI-RADS categories reflect increasing levels of radiographic suspicion and are linked to standardized management recommendations: category 0 denotes an incomplete assessment requiring additional imaging or clinical information; category 1 indicates no evidence of recurrence; category 2 reflects low suspicion, often favoring post-treatment change but warranting short-interval follow-up or direct inspection; category 3 indicates high suspicion for recurrence and typically prompts biopsy; and category 4 represents pathologically confirmed recurrence. Beyond creating a common reporting language, NI-RADS supports clinical decision-making by translating radiographic findings into actionable surveillance pathways. In contemporary head and neck oncology practice, pNI-RADS and nNI-RADS have become increasingly incorporated into follow-up imaging workflows and are recognized as clinically meaningful markers of recurrence risk and surveillance intensity^6–8^.

Comprehensive post-treatment surveillance, however, extends beyond NI-RADS category assignment alone. Clinical interpretation of surveillance imaging also depends on structured characterization of disease status, including the presence or absence of residual or recurrent primary tumor, evidence of nodal disease, and development of distant metastatic spread, as well as descriptive features such as primary site involvement, laterality, lesion size, nodal burden, and metastatic location. Together, these variables define the longitudinal disease state of OPC survivors and represent important inputs for recurrence tracking, risk modeling, clinical dashboarding, quality improvement initiatives, and survivorship research^9^. Capturing these surveillance descriptors in structured form at scale could substantially improve longitudinal monitoring of disease evolution and support downstream analytic applications across large survivorship cohorts.

Despite the growing importance of surveillance imaging data, NI-RADS categories and related disease-status descriptors are inconsistently captured as structured elements in routine clinical workflows and are most often embedded within free-text radiology narratives. Even when standardized reporting frameworks are available, substantial variation remains in reporting language, formatting, level of detail, and terminology used across radiologists and imaging modalities, creating challenges for systematic extraction and large-scale analysis^10^. Manual chart abstraction remains the most reliable approach for converting these narrative reports into analyzable structured variables, but this process is labor-intensive, costly, and impractical for longitudinal surveillance studies spanning hundreds or thousands of imaging examinations. Longitudinal survivorship cohorts may accumulate thousands of surveillance imaging studies over many years of follow-up, making comprehensive manual abstraction increasingly difficult to sustain^9^. These limitations create a major barrier to scalable imaging-based survivorship research and operational surveillance tracking.

Recent advances in large language models (LLMs) provide a promising solution for transforming unstructured clinical text into structured, reproducible data outputs. Unlike extraction of explicitly reported findings, NI-RADS assignment requires interpretation of radiologic descriptions and inference of recurrence risk categories based on post-treatment imaging features, creating a more clinically nuanced information extraction task. Prior work has demonstrated that LLM-based methods can accurately extract oncologic imaging findings and clinically meaningful concepts from narrative reports with strong agreement to expert review^11,12^. When deployed within enterprise-scale clinical data environments such as Palantir Foundry platform, LLM pipelines offer additional operational advantages, including secure handling of protected health information, schema-constrained outputs, prompt version control, scalable batch inference, integrated reviewer validation workflows, and transparent auditability of extraction logic^13^. These capabilities make enterprise LLM platforms particularly well suited for clinical surveillance extraction tasks that require both scalability and rigorous validation.

Given the clinical importance of post-treatment surveillance imaging in OPC survivorship and the limitations of free-text reporting for structured longitudinal analysis, there is a clear need for validated, high-throughput methods to reliably extract NI-RADS scores and key disease-status variables from radiology reports. In this study, we developed and validated a Foundry-based LLM extraction pipeline to infer pNI-RADS and nNI-RADS categories from narrative radiology reports and to extract structured surveillance variables describing primary tumor status, nodal disease, and distant metastases. Using a multi-reviewer adjudicated reference standard, we evaluated model agreement with expert review across both NI-RADS stratification and surveillance disease-status assessment, with the goal of enabling scalable structured imaging capture for longitudinal survivorship monitoring and downstream head and neck oncology research.

## Patients and Methods

### Inclusion Criteria

The analytic cohort included previously untreated patients with oropharyngeal cancer enrolled in the Stiefel MD Anderson Oropharyngeal Cancer (MDA-OPC) Cohort, a prospective observational cohort (PA14-0497) established through Core B of the OPC-SURVIVOR Program to support cross-project imaging, clinical, and survivorship research^9^. A local therapy completion date was defined for each patient as the latest recorded radiation therapy end date or surgical date, representing the end of curative-intent treatment. Imaging studies were linked to eligible patients using the image order dataset within Palantir Foundry, and only post-treatment examinations were retained, with all imaging performed prior to the therapy completion date excluded to remove baseline diagnostic studies.

From the linked dataset, we included only diagnostic head and neck imaging studies with available radiology impression text. Examinations not intended for post-treatment surveillance evaluation were excluded, including treatment planning or simulation studies, intraoperative imaging (OSI), interventional radiology ultrasound-guided procedures, and non-specific ultrasound studies. Reports lacking a clinically interpretable impression were also excluded.

### Baseline Characteristics

Baseline demographic and clinical characteristics were obtained by linking LLM-extracted imaging records to structured institutional datasets using medical record numbers (MRNs). Demographic variables, including age, sex, race, and ethnicity, were retrieved from the patient registry, while clinical information, including primary tumor subsite and stage, was derived from diagnosis and staging data after restricting to oropharyngeal cancer. Treatment information was obtained from structured treatment records, and multiple treatment entries per patient were consolidated into composite categories.

### Data Extraction and Model Validation

Structured imaging variables were extracted from unstructured radiology report text using a large language model pipeline implemented in Palantir Foundry (GPT 5.0). Two text fields from the institutional imaging dataset were used as inputs: narrative text and impression text. To focus the evaluation on reports requiring true language interpretation rather than direct label capture, imaging records with explicitly documented pNI-RADS or nNI-RADS scores were excluded. From this eligible set, 200 post-treatment imaging studies were randomly selected for LLM extraction and validation. Extraction workflows were built in Foundry Pipeline Builder using structured prompts, schema-constrained outputs, and stored supporting text for downstream review.

The LLM extracted two groups of variables. For NI-RADS assessment, the model used narrative and impression text to assign primary and nodal NI-RADS categories based on radiologic descriptions of post-treatment findings and recurrence risk, following the predefined study prompt and manual review framework; these outputs were inference-based rather than direct extraction of explicitly reported scores. From the narrative text, the model additionally extracted non–NI-RADS disease features, including primary tumor presence, site, laterality, and size; nodal disease presence, laterality, count, and largest nodal size; and presence and location of distant metastases, with supporting text captured for each variable.

Model performance was evaluated against a reference dataset curated through a structured three-reviewer manual review process in Fusion Sheet. Two reviewers independently annotated all sampled reports using standardized instructions for NI-RADS, primary tumor, nodal disease, and distant metastasis variables, with discrepancies adjudicated by a third reviewer to establish the final reference standard. LLM outputs were compared with adjudicated labels using agreement and F1-based metrics.

For NI-RADS evaluation, performance was assessed separately for low-risk (NI-RADS 1) and higher-risk categories (NI-RADS ≥2), with F1 score used for NI-RADS 1 and macro-F1 used to summarize performance across higher-risk ordinal categories. Additional measures included sensitivity, specificity, positive predictive value (PPV), and negative predictive value (NPV), with quadratic weighted kappa used to assess agreement across ordered NI-RADS categories beyond chance while accounting for the magnitude of disagreement between adjacent categories. For surveillance disease status variables, including primary tumor presence, nodal disease presence, and distant metastasis, performance was evaluated as binary classification tasks using agreement, F1 score, sensitivity, specificity, PPV, NPV, and Cohen’s kappa. Indeterminate or nonstandard labels were retained for descriptive reporting but excluded from binary metric calculations.

## Results

A total of 173 patients were included in the analysis. The median age at evaluation was 66.0 years (IQR, 60.0–73.0). The cohort was predominantly male (90.2%) and White (93.1%), with smaller proportions of Asian (2.3%) and Black or African American (2.9%) patients; 4.0% identified as Hispanic or Latino.

The primary tumor subsite was most commonly base of tongue (75 patients, 43.4%) and tonsil (74 patients, 42.8%), followed by oropharynx not otherwise specified (19 patients, 11.0%), soft palate (3 patients, 1.7%), and pharyngeal wall (2 patients, 1.2%). No primary subsite information was missing. Most patients presented with early-stage disease, including stage I (87 patients, 50.3%) and stage II (44 patients, 25.4%), while stage III and stage IV disease accounted for 28 patients (16.2%) and 14 patients (8.1%), respectively.

**Figure 1.**
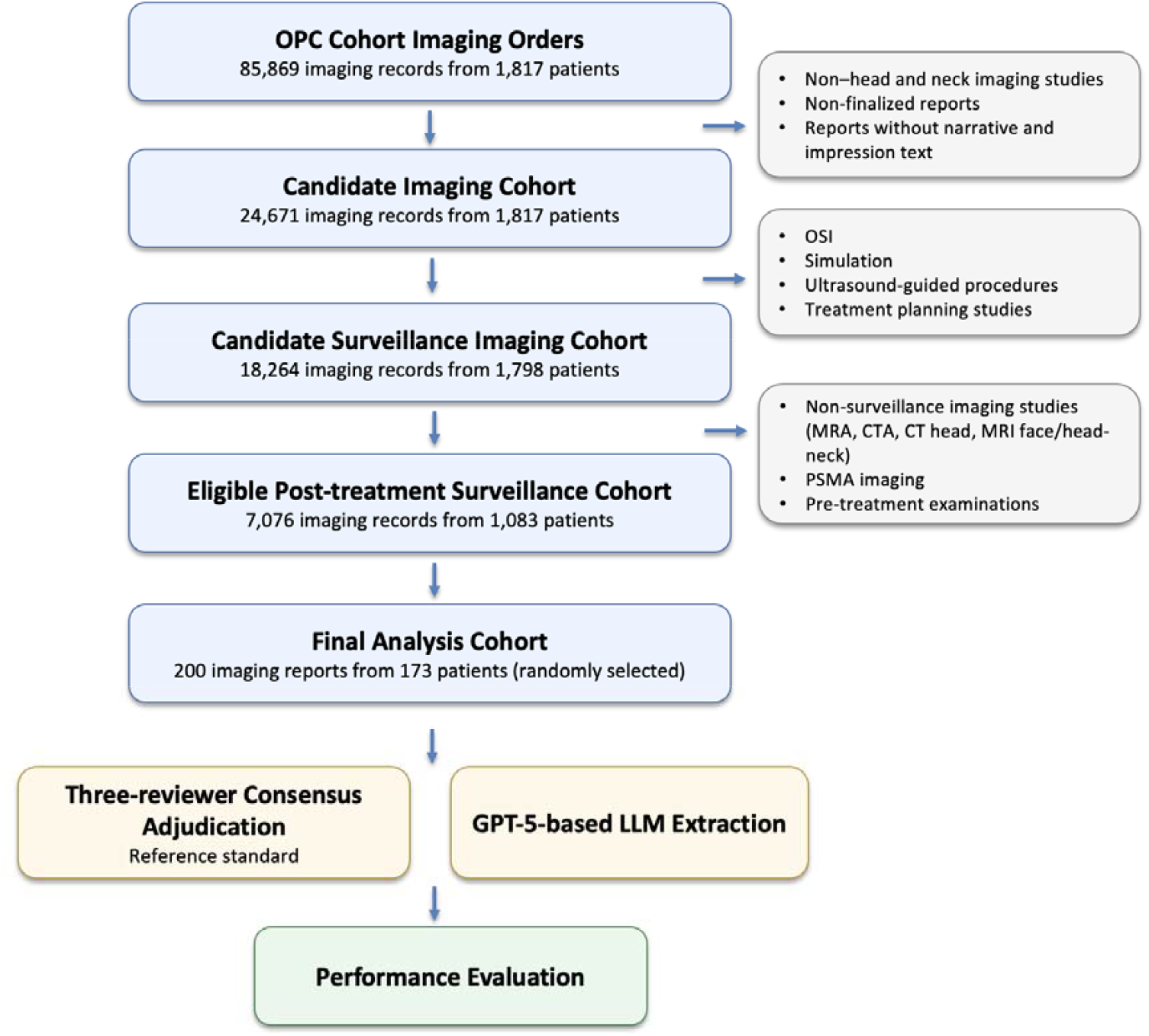
Cohort Selection and Validation Workflow for LLM–Based NI-RADS Extraction.

Regarding treatment, most patients received radiation-based therapy, including radiation with systemic therapy (113 patients, 65.3%) and radiation alone (30 patients, 17.3%). Smaller proportions received surgery with radiation and systemic therapy (16 patients, 9.2%), surgery alone (7 patients, 4.0%), surgery with radiation (4 patients, 2.3%), or systemic therapy alone (3 patients, 1.7%).

**Table 1.** Baseline characteristics of the study cohort.

| Characteristic | Value |
| --- | --- |
| Age, years | 66.0 (60.0, 73.0) |
| **Sex** |  |
| Male | 156 (90.2%) |
| Female | 17 (9.8%) |
| **Race** |  |
| White Or Caucasian | 161 (93.1%) |
| Black Or African American | 5 (2.9%) |
| Asian | 4 (2.3%) |
| American Indian Or Alaska Native | 1 (0.6%) |
| Other | 2 (1.2%) |
| **Ethnicity** |  |
| Not Hispanic Or Latino | 163 (94.2%) |
| Hispanic Or Latino | 7 (4.0%) |
| Unknown | 3 (1.7%) |
| **Primary subsite** |  |
| Base of tongue | 75 (43.4%) |
| Tonsil | 74 (42.8%) |
| Oropharynx (NOS) | 19 (11.0%) |
| Soft palate | 3 (1.7%) |
| Pharyngeal wall | 2 (1.2%) |
| **Stage group** |  |
| Stage I | 87 (50.3%) |
| Stage II | 44 (25.4%) |
| Stage III | 28 (16.2%) |
| Stage IV | 14 (8.1%) |
| **Treatment** |  |
| Radiation + systemic therapy | 113 (65.3%) |
| Radiation alone | 30 (17.3%) |
| Surgery + radiation + systemic therapy | 16 (9.2%) |
| Surgery alone | 7 (4.0%) |
| Surgery + radiation | 4 (2.3%) |
| Systemic therapy alone | 3 (1.7%) |

Values are median (IQR) for continuous variables and n (%) for categorical variables. Percentages were calculated using the full analytic cohort denominator unless otherwise indicated. Abbreviation: IQR, interquartile range.

**Table 2.** Distribution of NI-RADS scores assigned by reviewers and the LLM.

| NI-RADS domain | Score category | LLM | Reviewer |
| --- | --- | --- | --- |
| **Primary tumor site** |  |  |  |
|  | 1 | 135 (67.5%) | 143 (71.5%) |
|  | 2 | 39 (19.5%) | 32 (16.0%) |
|  | 3 | 9 (4.5%) | 11 (5.5%) |
|  | 4 | 7 (3.5%) | 10 (5.0%) |
|  | Indeterminate | 10 (5.0%) | 4 (2.0%) |
| **Nodal disease** |  |  |  |
|  | 1 | 142 (71.0%) | 151 (75.5%) |
|  | 2 | 38 (19.0%) | 30 (15.0%) |
|  | 3 | 8 (4.0%) | 5 (2.5%) |
|  | 4 | 3 (1.5%) | 8 (4.0%) |
|  | Indeterminate | 9 (4.5%) | 6 (3.0%) |

Values are n (%). Reviewer labels represent the final adjudicated reference standard. Indeterminate includes missing, blank, unknown, nonstandard, or non-1 to 4 labels. Abbreviations: LLM, large language model; NI-RADS, Neck Imaging Reporting and Data System.

**Table 3.** Agreement and classification performance for LLM-inferred NI-RADS scores.

| NI-RADS domain | Score category | Agreement, n/N (%) | F1 / Macro-F1 | Sensitivity | Specificity | PPV | NPV |
| --- | --- | --- | --- | --- | --- | --- | --- |
| **Primary tumor site** |  |  |  |  |  |  |  |
|  | NI-RADS 1 | 126/135 (93.3%) | 0.94 | 0.93 | 0.88 | 0.95 | 0.84 |
|  | NI-RADS 2 | 25/31 (80.6%) | 0.71 | 0.81 | 0.91 | 0.64 | 0.96 |
|  | NI-RADS 3 | 6/11 (54.5%) | 0.60 | 0.55 | 0.98 | 0.67 | 0.97 |
|  | NI-RADS 4 | 7/10 (70.0%) | 0.82 | 0.70 | 1.00 | 1.00 | 0.98 |
| | NI-RADS $\geq 2$ | 38/52 (73.1%) | 0.75 | 0.88 | 0.93 | 0.84 | 0.95 |
| **Nodal disease** |  |  |  |  |  |  |  |
|  | NI-RADS 1 | 130/144 (90.3%) | 0.93 | 0.90 | 0.83 | 0.95 | 0.71 |
|  | NI-RADS 2 | 22/30 (73.3%) | 0.65 | 0.73 | 0.90 | 0.58 | 0.95 |
|  | NI-RADS 3 | 2/4 (50.0%) | 0.33 | 0.50 | 0.97 | 0.25 | 0.99 |
|  | NI-RADS 4 | 3/8 (37.5%) | 0.55 | 0.38 | 1.00 | 1.00 | 0.97 |
| | NI-RADS $\geq 2$ | 27/42 (64.3%) | 0.56 | 0.83 | 0.90 | 0.71 | 0.95 |

Reviewer-adjudicated labels were used as the reference standard. For NI-RADS 1 to 4, F1 reflects one-vs-all classification. For NI-RADS $\geq 2$ , macro-F1 reflects exact category agreement across reviewer-labeled NI-RADS 2 to 4 cases. Weighted $\kappa$ used quadratic weighting. Indeterminate labels were excluded from numeric performance calculations. Abbreviations: LLM, large language model; NI-RADS, Neck Imaging Reporting and Data System; PPV, positive predictive value; NPV, negative predictive value.

For primary tumor site classification, agreement between LLM-inferred and adjudicated reviewer-assigned NI-RADS scores was 93.3% (126/135) for pNI-RADS 1, with an F1 score of 0.94. For higher-risk primary categories (pNI-RADS ≥2), exact category agreement was 73.1% (38/52), with a macro-F1 of 0.75, sensitivity of 0.88, specificity of 0.93, PPV of 0.84, and NPV of 0.95. Performance across individual higher-risk primary categories remained favorable, with strongest classification observed for pNI-RADS 4. Overall ordinal agreement was high, with quadratic weighted κ values of 0.90 for pNI-RADS and 0.79 for nNI-RADS. Among reviewer-labeled NI-RADS ≥2 examinations, weighted κ values were 0.81 for pNI-RADS and 0.59 for nNI-RADS.For nodal disease classification, agreement was 90.3% (130/144) for nNI-RADS 1, with an F1 score of 0.93. For higher-risk nodal categories (nNI-RADS ≥2), exact category agreement was 64.3% (27/42), with a macro-F1 of 0.56, sensitivity of 0.83, specificity of 0.90, PPV of 0.71, and NPV of 0.95. Among individual higher-risk nodal categories, classification was most consistent for nNI-RADS 2, while performance for nNI-RADS 3 and 4 was more variable. Quadratic weighted kappa for nodal classification was 0.59.

**Table 4.** Distribution of disease-status labels assigned by reviewers and the LLM.

| Disease-status variable | Status category | LLM | Reviewer |
| --- | --- | --- | --- |
| **Primary tumor present** |  |  |  |
|  | Present | 29 (14.5%) | 44 (22.0%) |
|  | Absent | 137 (68.5%) | 139 (69.5%) |
|  | Indeterminate | 34 (17.0%) | 17 (8.5%) |
| **Nodal disease present** |  |  |  |
|  | Present | 71 (35.5%) | 83 (41.5%) |
|  | Absent | 118 (59.0%) | 111 (55.5%) |
|  | Indeterminate | 11 (5.5%) | 6 (3.0%) |
| **Distant metastasis present** |  |  |  |
|  | Present | 15 (7.5%) | 11 (5.5%) |
|  | Absent | 143 (71.5%) | 150 (75.0%) |
|  | Indeterminate | 42 (21.0%) | 39 (19.5%) |

Values are n (%). Reviewer labels represent the final adjudicated reference standard. Indeterminate includes missing, blank, unknown, or nonstandard labels. Abbreviation: LLM, large language model.

**Table 5.**
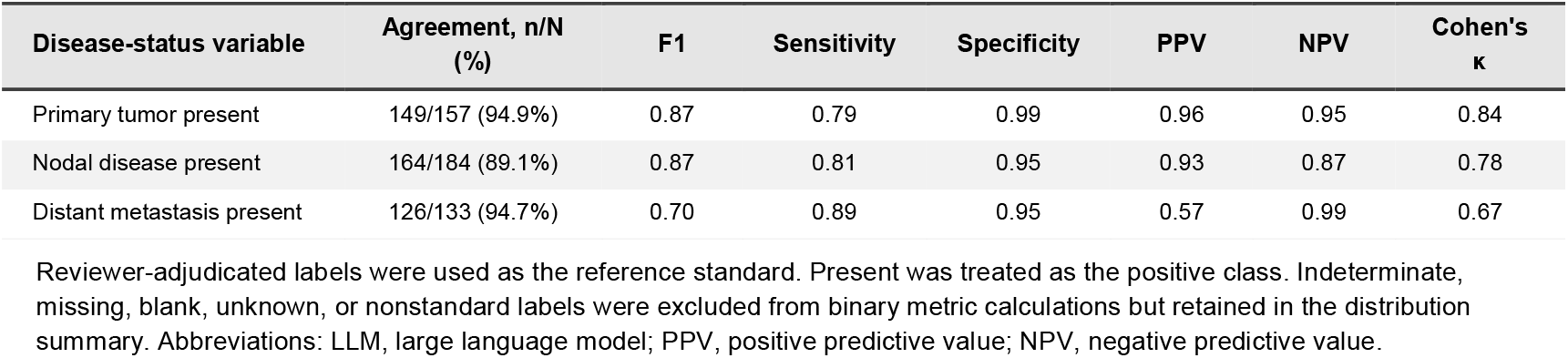
Agreement and classification performance for LLM-inferred disease-status variables.

| Disease-status variable | Agreement, n/N (%) | F1 | Sensitivity | Specificity | PPV | NPV | Cohen's $\kappa$ |
| --- | --- | --- | --- | --- | --- | --- | --- |
| Primary tumor present | 149/157 (94.9%) | 0.87 | 0.79 | 0.99 | 0.96 | 0.95 | 0.84 |
| Nodal disease present | 164/184 (89.1%) | 0.87 | 0.81 | 0.95 | 0.93 | 0.87 | 0.78 |
| Distant metastasis present | 126/133 (94.7%) | 0.70 | 0.89 | 0.95 | 0.57 | 0.99 | 0.67 |

Reviewer-adjudicated labels were used as the reference standard. Present was treated as the positive class. Indeterminate, missing, blank, unknown, or nonstandard labels were excluded from binary metric calculations but retained in the distribution summary. Abbreviations: LLM, large language model; PPV, positive predictive value; NPV, negative predictive value.

For post-treatment disease surveillance assessment, agreement between LLM-inferred and adjudicated reviewer-assigned labels was high across primary tumor presence, nodal disease presence, and distant metastasis variables. Agreement for primary tumor presence or absence was 94.9% (149/157), with an F1 score of 0.87, sensitivity of 0.79, specificity of 0.99, PPV of 0.96, NPV of 0.95, and Cohen’s κ of 0.84. Agreement for nodal disease presence or absence was 89.1% (164/184), with an F1 score of 0.87, sensitivity of 0.81, specificity of 0.95, PPV of 0.93, NPV of 0.87, and κ of 0.78. Agreement for distant metastasis presence or absence was 94.7% (126/133), with an F1 score of 0.70, sensitivity of 0.89, specificity of 0.95, PPV of 0.57, NPV of 0.99, and κ of 0.67. LLM-derived label distributions were generally similar to reviewer adjudication, although indeterminate assignments were modestly more frequent in LLM outputs, particularly for primary tumor status.

## Discussion

In this study, we developed and validated an LLM-based extraction pipeline within Palantir Foundry to derive NI-RADS classifications and post-treatment disease-status variables from unstructured radiology reports in OPC survivors. Using a three-reviewer adjudicated reference standard, the LLM demonstrated strong agreement with expert review for both primary and nodal NI-RADS assessment, particularly for low-risk surveillance examinations. The model also achieved high performance in identifying clinically relevant disease-status variables, including primary tumor recurrence, nodal disease, and distant metastases. Together, these findings demonstrate the feasibility of converting narrative surveillance imaging reports into structured longitudinal data while maintaining strong concordance with expert interpretation.

A notable finding was the consistently strong performance observed for surveillance disease-status variables. Agreement approached 90% to 95% for primary tumor, nodal disease, and distant metastasis, with high specificity and NPV across all evaluated domains. Given that most surveillance imaging examinations in OPC survivorship do not demonstrate recurrent disease, accurate identification of negative examinations is particularly important for longitudinal monitoring and cohort construction. These results suggest that LLM-derived disease-status variables can serve as reliable structured inputs for recurrence tracking, clinical dashboards, survivorship registries, and imaging-based outcomes research^9^.

Our findings are consistent with a growing body of literature demonstrating the utility of LLMs for extracting clinically meaningful information from radiology reports. Prior studies have shown strong performance for extraction of imaging recommendations, structured diagnostic findings, and disease-specific radiologic features from free-text reports^11,12,14^. However, most existing work has focused on extraction of explicitly stated findings. In contrast, NI-RADS classification requires reasoning of post-treatment imaging descriptions and assignment of structured risk categories that reflect radiologic suspicion for recurrence. This represents a more clinically nuanced task that more closely resembles expert radiologic reasoning. To our knowledge, this is among the first studies to evaluate LLM-based extraction of NI-RADS classifications from post-treatment head and neck surveillance imaging reports. The favorable performance observed in this study suggests that modern LLMs are capable of operationalizing standardized surveillance frameworks from heterogeneous narrative reports and may facilitate scalable capture of imaging-derived risk information in routine clinical practice.

An additional strength of this work is the implementation of the extraction workflow within the Foundry environment^15^. Beyond model performance alone, enterprise-scale clinical data platforms provide important operational advantages, including secure handling of protected health information, reproducible prompt management, schema-constrained outputs, integrated reviewer validation workflows, and transparent auditability of extraction logic. These features support deployment of LLM-based extraction pipelines within large clinical research programs and create opportunities for longitudinal integration of imaging, clinical, and patient-reported outcome data across survivorship cohorts.

Although overall performance was favorable, agreement was lower for higher-risk NI-RADS categories, particularly for nodal classifications. Several factors likely contributed to this finding. First, higher-risk surveillance findings were uncommon within the validation cohort, with relatively few NI-RADS 3 and NI-RADS 4 examinations available for evaluation. Under these circumstances, even a small number of disagreements can substantially influence performance metrics, resulting in greater variability in estimates of agreement, sensitivity, and F1 scores. Second, interpretation of suspicious post-treatment findings often depends on subtle radiologic descriptors and contextual language that may vary across reports and reviewers. Finally, some disagreement likely reflects inherent variability in human interpretation of post-treatment imaging rather than purely model-related error^16^. Despite these challenges, weighted κ values demonstrated moderate to substantial agreement across ordered NI-RADS categories, supporting the overall reliability of the extraction framework.

Several limitations should be considered when interpreting these findings. This study was conducted at a single tertiary cancer center using a specific OPC survivorship population, which may limit generalizability to other institutions, reporting styles, and patient populations. Although rigorous three-reviewer adjudication was used to establish the reference standard, manual review is inherently labor-intensive and represents a major barrier to validating large-scale imaging datasets. Consequently, validation was performed on a randomly selected sample of 200 imaging reports rather than the full surveillance cohort. This limitation highlights one of the primary motivations for developing automated extraction methods, as manual abstraction of thousands of longitudinal imaging examinations is often impractical in survivorship research. In addition, because higher-risk NI-RADS categories were relatively rare, larger validation cohorts and longer-term surveillance follow-up will be important to obtain more precise estimates of agreement for uncommon but clinically significant findings^8,16^. Future studies incorporating larger multi-institutional cohorts and external validation datasets will be essential to further establish the generalizability and robustness of LLM-based surveillance extraction pipelines.

Collectively, these findings demonstrate that LLM-based extraction can provide an effective and scalable approach for transforming unstructured radiology reports into structured and timely surveillance data. As survivorship cohorts continue to expand and longitudinal imaging data become increasingly important for clinical monitoring and research, automated extraction frameworks to alert clinicians and research teams of disease status changes may facilitate near real-time clinical decision-making and substantially reduce abstraction burden while enabling large-scale analysis of imaging-derived outcomes across head and neck oncology populations^9,17–19^.

## Conclusion

A Foundry-based LLM extraction pipeline demonstrated strong agreement with expert review for automated derivation of NI-RADS classifications and key post-treatment disease-status variables from unstructured radiology reports in OPC survivors. The approach enabled reliable conversion of narrative surveillance imaging into structured longitudinal data while substantially reducing the need for manual abstraction. These findings support the use of enterprise-scale LLM workflows to facilitate imaging-based survivorship monitoring, clinical analytics, and large-scale head and neck oncology research. Future multi-institutional validation efforts will help further define the role of LLM-based extraction systems in supporting standardized surveillance data capture across diverse clinical settings.

## Data Availability

Anonymized and de-identified data that support the findings of this study are openly available in Figshare at https://doi.org/10.6084/m9.figshare.32620149.

## Pre-print availability statement

In accordance with NIH Policy NOT-OD-17-050, *Reporting Preprints and Other Interim Research Products*, which specifies: “The NIH encourages investigators to use interim research products, such as preprints, to speed the dissemination and enhance the rigor of their work”, we have deposited a pre-peer review version of the initial version of this submitted manuscript at medrxiv with DOI available upon acceptance.

## Data sharing statement

In accordance with NOT-OD-21-013, Final NIH Policy for Data Management and Sharing, anonymized/de-identified data that support the findings of this study are openly available in an NIH-supported generalist scientific data repository (FigShare) at 10.6084/m9.figshare.32620149.

## Public access policy compliance

This study was approved by the Institutional Review Board of The University of Texas MD Anderson Cancer Center. Patient data were derived from the Oropharynx Cancer Registry Database protocol (PA14-0947), whereby patients consent to general use of their data for research, and analyzed under protocol 2024-0002.

## Reporting Guideline Compliance

Study design, model evaluation, and reporting were informed by the Framework for LLM Assessment in Radiology (FLAIR) reporting recommendations where applicable.

## Contributor Role Taxonomy (CRediT) Statement

**Wenye Song:** Conceptualization, Data curation, Formal analysis, Investigation, Methodology, Validation, Visualization, Writing – original draft, Writing – review & editing.

**Lina Shbita:** Validation, Writing – review & editing.

**Isabelle Jia Hui Jang:** Validation, Writing – review & editing.

**Olga Starostina:** Data curation, Resources, Software.

**Ryan Lewis:** Data curation, Resources, Software.

**Ariana J. Sahli:** Data curation.

**Warren Floyd:** Methodology, Resources, Validation.

**Md Mahin:** Methodology, Validation.

**Waree Rinsurongkawong:** Data curation.

**Carly E. A. Barbon:** Resources, Supervision, Writing – review & editing.

**Stephen Y. Lai:** Funding acquisition, Resources, Supervision.

**J. Jack Lee:** Formal analysis, Methodology, Supervision, Writing – review & editing.

**Komal Shah:** Investigation, Validation.

**Melissa Chen:** Writing – review & editing.

**Katherine A. Hutcheson:** Funding acquisition, Methodology, Project administration, Resources, Supervision, Writing – review & editing.

**Clifton David Fuller:** Conceptualization, Funding acquisition, Methodology, Resources, Supervision, Writing – review & editing.

**Amy C. Moreno:** Conceptualization, Funding acquisition, Methodology, Project administration, Resources, Software, Supervision, Validation, Writing – review & editing.

## Acknowledgements

This work was enabled by The University of Texas MD Anderson Cancer Center Context Engine and the Context Engine Team. The Context Engine is MD Anderson’s institutional Data Management System and Digital Architecture.

